# Rapid real-time tracking of non-pharmaceutical interventions and their association with SARS-CoV-2 positivity: The COVID-19 Pandemic Pulse Study

**DOI:** 10.1101/2020.07.29.20164665

**Authors:** Steven J. Clipman, Amy P. Wesolowski, Dustin G. Gibson, Smisha Agarwal, Anastasia S. Lambrou, Gregory D. Kirk, Alain B. Labrique, Shruti H. Mehta, Sunil S. Solomon

## Abstract

**Background:** Current mitigation strategies for severe acute respiratory syndrome coronavirus 2 (SARS-CoV-2) rely on population-wide adoption of non-pharmaceutical interventions (NPIs). Collecting demographically and geographically resolved data on NPIs and their association with SARS-CoV-2 infection history can provide critical information related to reopening geographies.

**Methods:** We sampled 1,030 individuals in Maryland from June 17 – June 28, 2020 to capture socio-demographically and geographically resolved information about NPI adoption, access to SARS-CoV-2 testing, and examine associations with self-reported SARS-CoV-2 positivity.

**Results:** Median age of the sample was 43 years and 45% were men; Whites and Blacks/African Americans represented 60% and 23%, respectively. Overall, 96% of the sample reported traveling outside their home for non-employment related services: most commonly cited reasons were essential services (92%) and visiting friends/family (66%). Use of public transport was reported by 18% of respondents. 68% reported always social distancing indoors and 53% always wearing masks indoors; indoor social distancing was significantly less common among younger vs. older individuals, and race/ethnicity and income were significantly associated with mask use (p<0.05 for all). 55 participants (5.3%) self-reported ever testing positive for SARS-CoV-2 with strong dose-response relationships between movement frequency and SARS-CoV-2 positivity that were significantly attenuated by social distancing. In multivariable analysis, history of SARS-CoV-2 infection was negatively associated with the practice of social distancing (adjusted Odd Ratio [aOR]: 0.10; 95% Confidence Interval: 0.03 – 0.33); the only travel associated with higher likelihood of SARS-CoV-2 infection was use of public transport (aOR for ≥7 times vs. never: 4.29) and visiting a place of worship (aOR for ≥3 times vs. never: 16.0) after adjusting for social distancing.

**Conclusions:** Using a rapid cost-efficient approach, we highlight the role of movement and social distancing on SARS-CoV-2 transmission risk. Continued monitoring of NPI uptake, access to testing, and the subsequent impact on SARS-CoV-2 transmission will be critical for pandemic control and decisions about reopening geographies.

**Key Points:** *What we did:*

- We utilized an online survey approach to sample residents of Maryland consistent with the distributions of age, gender, race/ethnicity, and income in the state.
- We asked questions about places (and the frequency) visited for essential and nonessential services in the prior 2 weeks, practice of non-pharmaceutical interventions (NPIs) while visiting various places, and access to SARS-CoV-2 testing.
- We characterized how movement and adoption of NPIs differed by key demographics (age, race, gender, income) and how these were associated with self-reported SARS-CoV-2 positivity.

*What we found:*

- 96% of the sample reported traveling for either essential or non-essential services in the prior 2 weeks; 82% reported traveling for non-essential services.
- The adoption of NPIs varied by age, race/ethnicity, and income.
- Self-reported SARS-CoV-2 positivity was highest among Latinos followed by Blacks/African Americans and then Whites.
- The more frequently a person traveled/visited places for non-essential services, the more likely they were to report ever having tested positive for SARS-CoV-2.
- The strict practice of social distancing was associated with a lower likelihood of ever having tested positive for SARS-CoV-2; moreover, strict social distancing attenuated the association between most forms of movement and SARS CoV-2 positivity
- Using public transport and attending places of worship remained associated with a higher likelihood of having tested positive for SARS-CoV-2 even when practicing social distancing.
- About 70% of people who wanted a SARS-CoV-2 test were able to get a test but there were delays of a week or more from wanting a test to getting a result among the majority of the sample.

*What it means:*

- The more people move the more likely they are to test positive for SARS-CoV-2; if you must travel, practice social distancing as it reduces the likelihood of testing positive.
- Avoid public transport to the extent possible.
- Strategies to reduce time from wanting a test to getting a result are critical to enhance early case detection and isolation to curb transmission.

## Introduction

The severe acute respiratory syndrome coronavirus 2 (SARS CoV-2) and associated Coronavirus Disease 2019 (COVID-19) pandemic continues to evolve at a rapid pace, having affected more than 15 million persons globally and more than 4 million in the US as of July 23, 2020.^1^ While there has been rapid progress in therapeutic^2,3^ and vaccine development,^4,5^ nothing to date is a panacea. The primary means of curtailing community transmission remains testing and contact tracing, and continued implementation of non-pharmaceutical interventions (NPIs) such as social distancing and masking.^6,7^

Monitoring trends in the adoption of these NPIs may provide insight into the trajectory of local SARS-CoV-2 epidemics, information that can guide public health practice and policy.^8^ First, declining or poor levels of NPI adoption could signal an impending upsurge in cases indicating a need to mobilize resources to medical facilities and expand testing to facilitate early diagnosis and isolation. Second, these data could help to target messaging based on identification of population strata or geographic areas where NPI adherence is low. Third, high levels of NPI adoption could support decisions related to re-opening businesses. Given the constantly changing public health guidance around NPIs and potential NPI fatigue, jurisdictions should monitor these behaviors at frequent intervals.

Several reports have used geolocation data from mobile phone users as a surrogate for social distancing and mobility.^9-12^ These analyses, however, are typically ecological in nature and are not able to capture finer scale changes in behavior including identification of subgroups (e.g., race/ethnicity, income, age) who are differentially practicing social distancing. Further, mobile phone data cannot distinguish between travel for essential services/employment versus leisure. Others have used online symptom trackers and/or surveys to capture information on attitudes towards NPIs;^13-15^ however, most have not collected information on NPI adoption across the spectrum of different activities in which individuals engage.

We used an online panel to rapidly sample persons in Maryland to capture granular information on NPI adoption, travel, access to SARS-CoV-2 testing, and associations with self-reported SARS-CoV-2 positivity.

## Methods

### Study Setting

As of July 23, 2020, Maryland had nearly 81,000 confirmed COVID-19 cases, more than 3,200 deaths and a current positive test proportion of 4.56%.^16^ Community transmission has been sustained since March 12, 2020,^16^ and statewide stay-at-home orders were implemented from March 30 to May 15, 2020. As of April 18, 2020, Maryland required face coverings on public transit and within retail outlets for employees and customers. Stage Two of phased reopening began on June 12, 2020, allowing indoor restaurant dining to resume at 50% capacity; on June 19, 2020, gyms, retail stores, salons, barbershops, nail salons, amusement parks, outdoor pools, and indoor worship services were permitted to open with capacity restrictions.^17^

### Study Sample

Participants were recruited from across Maryland from June 17 – June 28, 2020 using Dynata (https://www.dynata.com), one of the largest first-party global data platforms. Dynata maintains a database of potential participants who are randomized to specific surveys if they meet the specified demographic targets of the survey; participants receive modest compensation for participation. Security checks and quality verifications are used to verify identity and prevent duplication before participants begin surveys. These include digital fingerprinting to prevent duplication, spot checking via third party verification to prove identity and a dedicated panelist quality team. Dynata staff are active in multiple online research quality initiatives including the European Society for Opinion and Marketing Research (ESOMAR).

In order to accrue a sample representative of Maryland, we provided Dynata with quotas for age, gender, race/ethnicity, and income based on the population composition of Maryland. Quotas were monitored through the Dynata internal router system and self-reported survey data. Dynata distributed survey invitations to 2,322 persons. Of these, 1,466 visited the survey site and responded to at least one survey question – 109 started but did not complete the survey and 310 responses were excluded for non-eligibility (age less than 18 years or current residence outside Maryland). We further excluded 17 participants who did not provide a response to ever being tested for SARS-CoV-2, for a final sample size of 1,030 (target = 1000).

### Survey

The electronic survey was based on a combination of existing COVID-19 surveys and new questions on multiple domains including sociodemographic characteristics, adoption of NPIs (including social distancing and mask wearing) and access to SARS-CoV-2 testing. All questions were asked with respect to the prior two weeks. To understand social distancing practices, participants were asked about the types and frequency with which they visited places/locations in the prior two weeks including indoor locations such as large gatherings, homes of friends and family members, gyms, salons, grocery stores, pharmacies, restaurants, places of worship, and outdoor locations such as beaches and pools. Participants were also asked about social distancing practices and mask wearing at these locations.

### Statistical Analysis

Chi square tests and Mann-Whitney tests were used to compare categorical and continuous variables, respectively. Logistic regression was used to identify variables associated with ever testing positive for SARS-CoV-2 infection. Variables of interest included demographics, mobility patterns including types of outdoor and indoor locations visited in the prior two weeks, and adoption of preventive behaviors such as masking and social distancing. Variables were included in the multivariable models if they held biological significance, were statistically significant in univariable models (p<0.05) and/or had significant variable importance scores as determined by machine learning feature selection using random forest with a Boruta wrapper algorithm.^18^ As a sensitivity analyses, we restricted the outcome to self-report of a recent (prior 2 weeks) SARS-CoV-2 positive test.

### Ethical Clearance

The study was approved by the Institutional Review Board of the Johns Hopkins Bloomberg School of Public Health (IRB00012413) and all participants consented to participate.

## Results

### Characteristics of study population

The median age of the 1,030 participants was 43 years and 55% were women. The majority (n=618 [60%]) were White; 239 (23%) and 74 (7%) self-identified as Black/African American and Hispanic/Latino, respectively (Table 1). About 69% of the participants reported attending college or possessing a graduate degree, and the majority (55%) reported a household income less than $70,000. There were 303 (29%) respondents who reported working outside of the home at the time of the survey. Participants were sampled from all 24 counties in Maryland with the largest representation from the most populous counties (Fig 1). In general, sociodemographic characteristics of the study sample were highly representative of the Maryland state population.

**Table 1.**
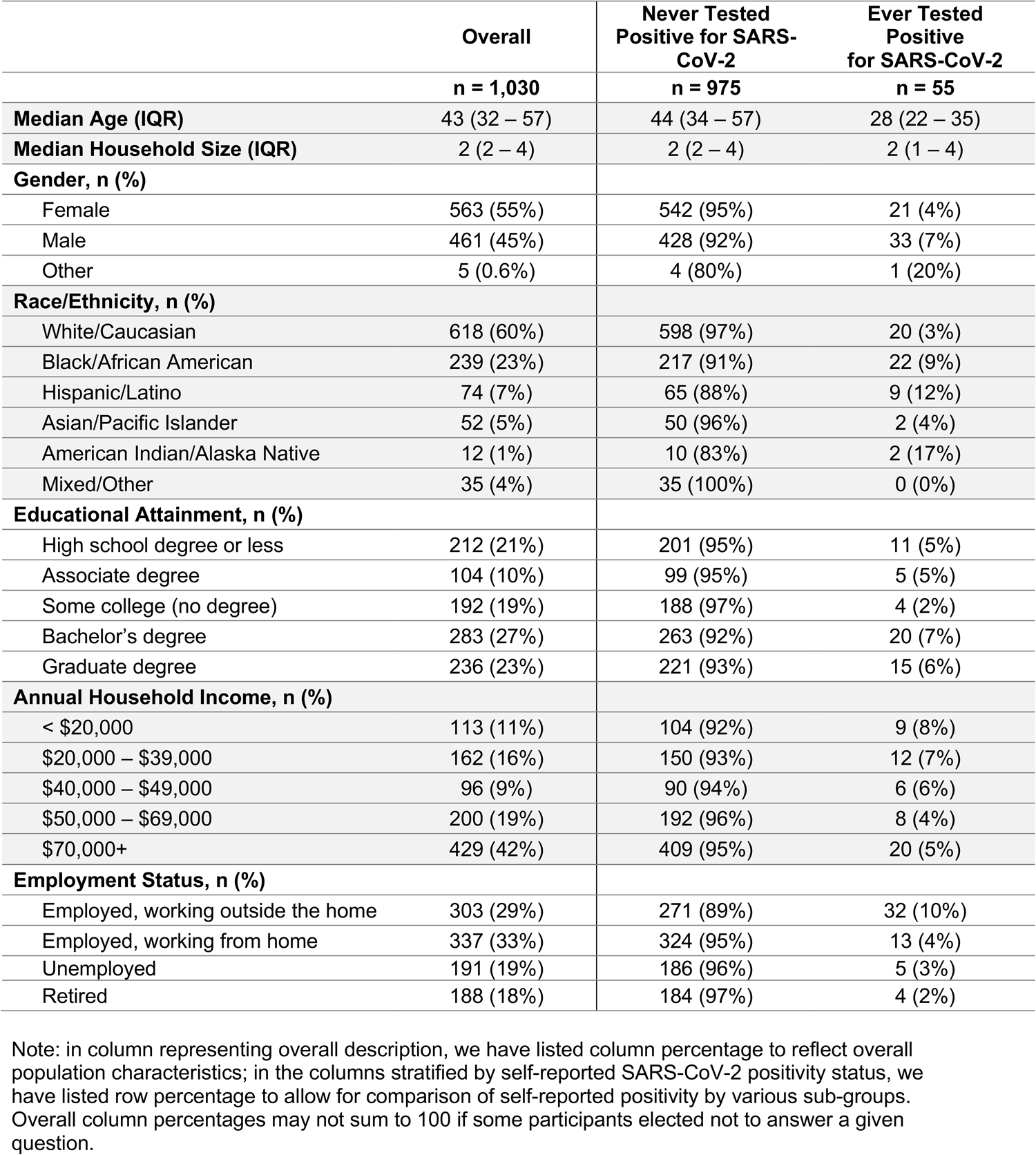
Characteristics of study sample by self-reported SARS-CoV-2 infection status

**Fig 1.**
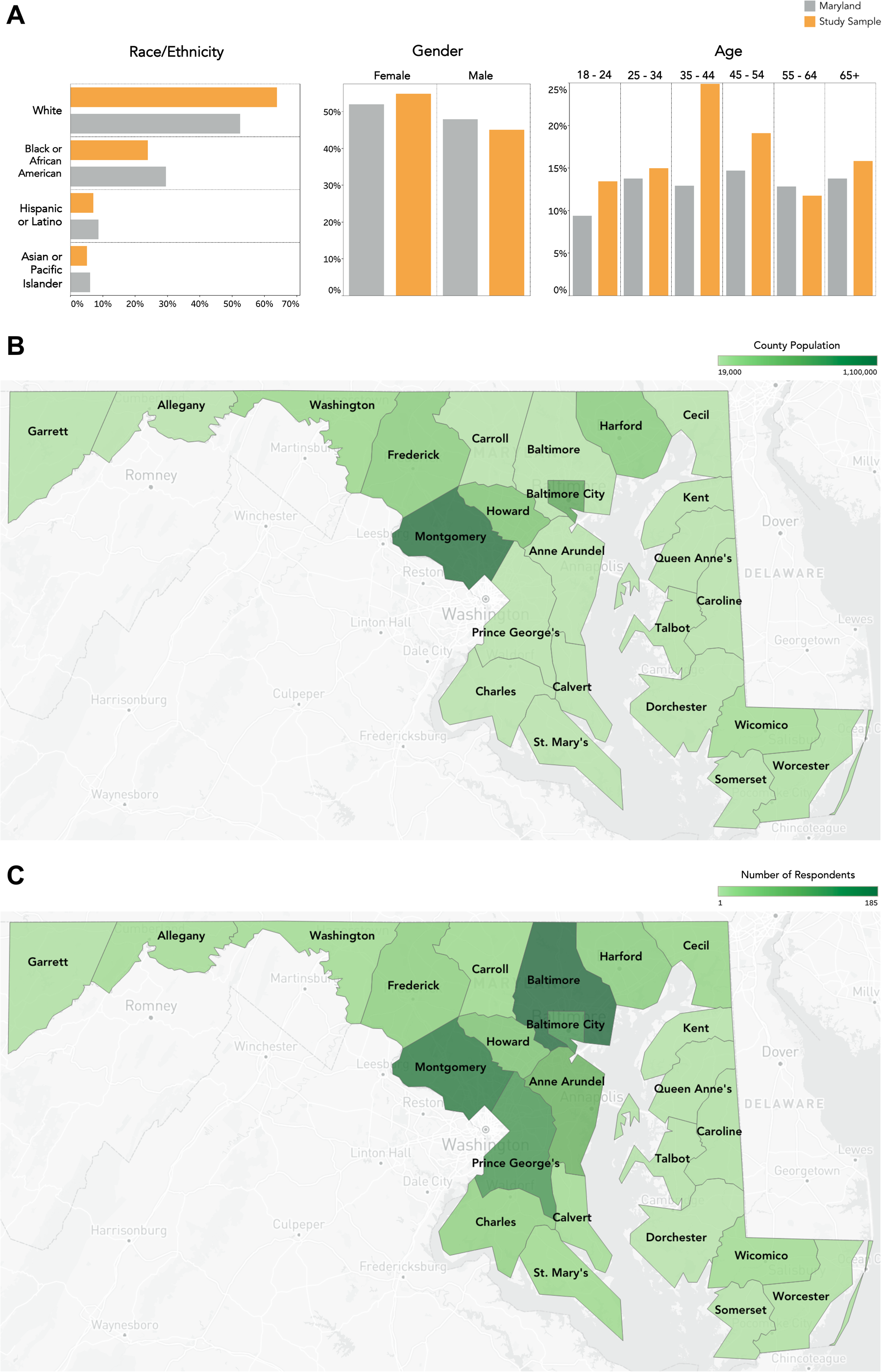
(A) Distribution of select population demographics in Maryland and the study sample, and (B) Maryland population by county compared to (C) number of survey responses by county. Source: United States Census Bureau. B01001 SEX BY AGE, 2018 American Community Survey 5-Year Estimates. U.S. Census Bureau, American Community Survey Office. Web. 19 December 2019. http://www.census.gov/. United States Census Bureau. Annual Estimates of the Resident Population: April 1, 2010 to July 1, 2019. U.S. Census Bureau, Population Division. Web. July 2020. http://www.census.gov/.

### Adoption of non-pharmaceutical interventions

Overall, 990 (96%) participants reported leaving their home at least once during the prior two weeks. Of these participants, almost all (92%) reported traveling for essential services (e.g., grocery store or pharmacy) at least once and 40% reported going 3 or more times in the prior two weeks. Following travel for essential services, the next most frequented places were visiting friends/family (66%), indoor venues such as bars, salons or restaurants (49%) and outdoor venues such as beaches or pools (25%) (Fig 2). Among those who reported travel, 18% and 15% reported using public transport and visiting a place of worship, respectively. There were 5% who reported engaging in all of these activities. About a quarter (26%) reported attending at least one gathering of 10 or more people in the prior two weeks, of whom 8% reported attending 3 or more; 104 respondents (10%) reported attending at least one gathering of 100 persons or more.

**Fig 2.**
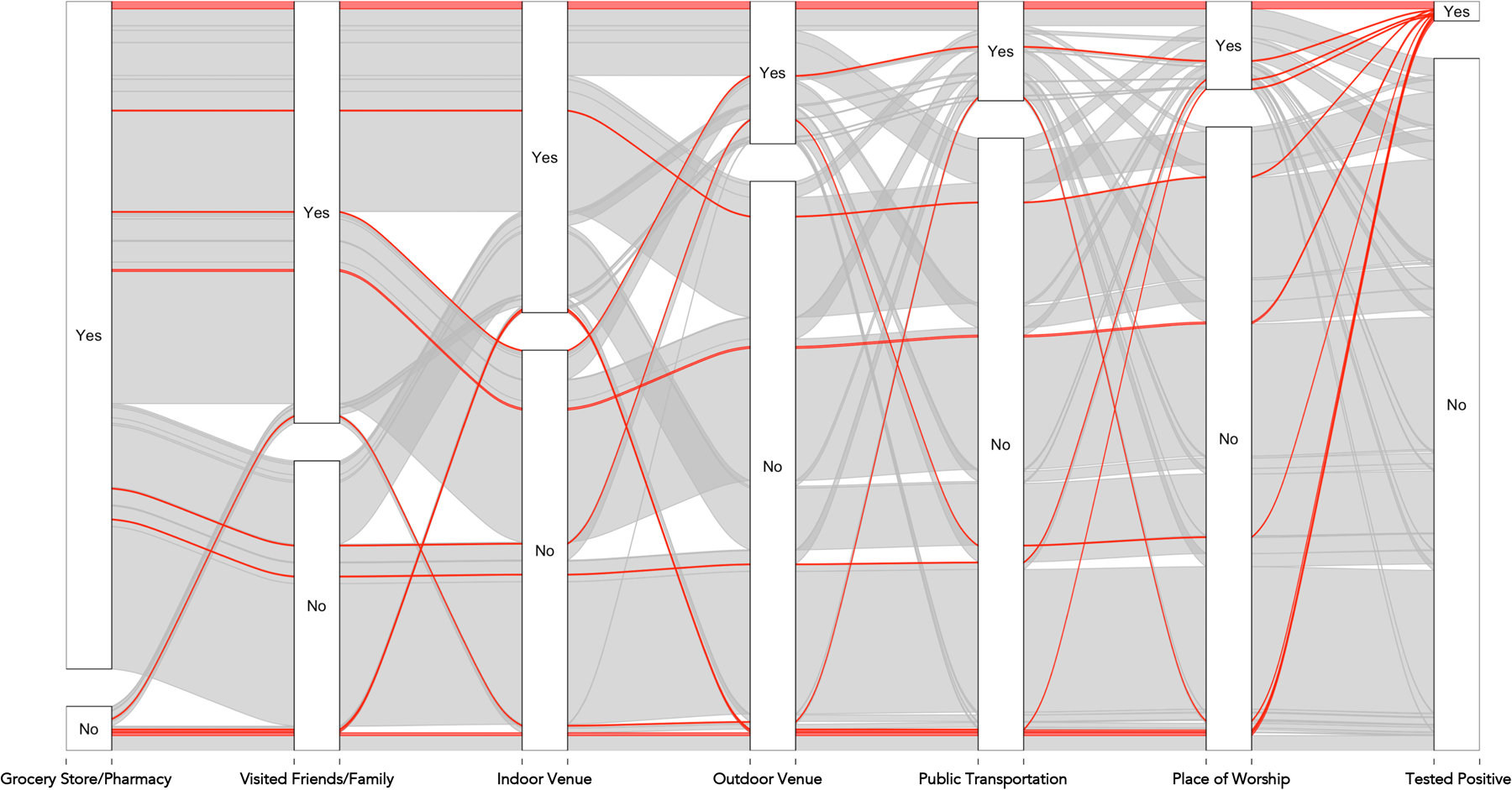
Sankey diagram showing participant responses across all questions capturing mobility in the past 2 weeks. Participant responses are depicted in each rectangular node with flows representing how many individuals have responded across the entire set of questions, for example, individuals who responded “yes” to both traveling to a grocery store/pharmacy and visiting friends/family are reflected in the flow connecting the two “yes” nodes for these variables. Individuals who have ever tested positive for SARS-CoV-2 are shown in red, whereas all others are shown in grey. A total of 51 (5%) participants responded “yes” to all mobility questions and 39 (4%) responded “no” to all; 17 (31%) of those who tested positive for SARS-CoV-2 responded “yes” to all and 2 (4%) responded “no” to all. The thickness of the flow is directly proportional to the number of respondents who report that behavior.

The majority reported practicing social distancing when visiting indoor and outdoor locations, although the reported adoption of social distancing increased with age. For example, 81% of those over the age of 65 reported always practicing social distancing at outdoor activities compared to 58% of those 18-24 years of age (p<0.001; Fig 3). About half (53%) reported that they always wore a mask when visiting indoor and outdoor locations; 17% and 19% reported never wearing a mask when visiting indoor or outdoor locations, respectively. While age was not significantly associated with self-reported mask use, race/ethnicity and income were. About three-fourths (72%) of Blacks reported always wearing a mask outdoors compared to 44% of Whites (p<0.001). Further, 62% of those earning household income less than $20,000/year reported always wearing a mask outdoors compared to 48% of those with household income of $70,000 or greater (p=0.018).

**Fig 3.**
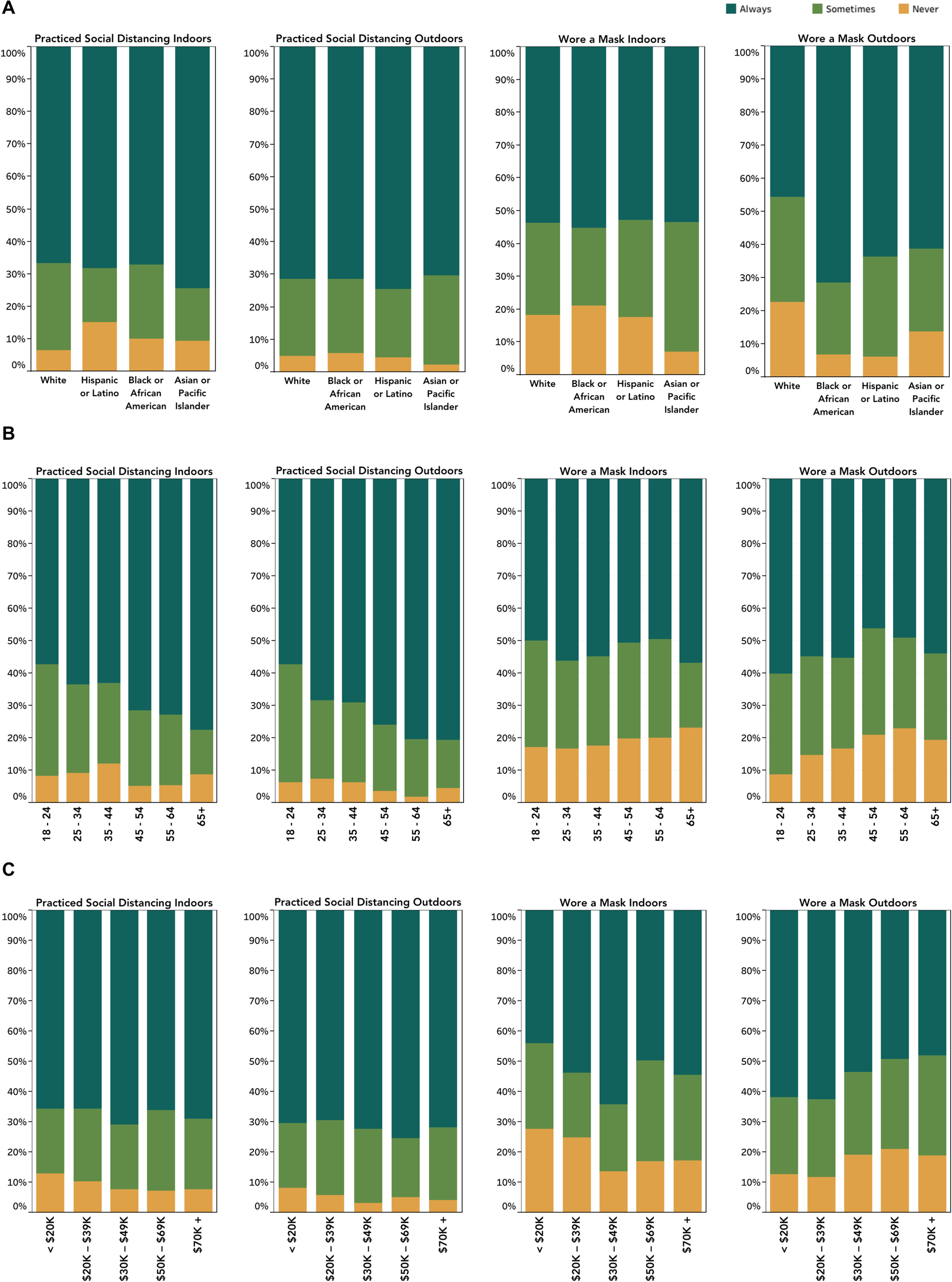
Self-reported uptake of non-pharmaceutical interventions in the prior 2 weeks by (A) race/ethnicity, (B) age, and (C) annual household income.

### SARS-CoV-2 Infection and Access to Testing

Overall, 55 participants (5.3%) self-reported ever testing positive for SARS-CoV-2 infection. Participants who self-reported having tested positive were significantly more likely to be male, younger, self-identify as Black/African American or Hispanic/Latino, and be required to travel outside of home for work (Table 1). The distribution of self-reported SARS-CoV-2 positivity by Maryland County is presented in Supplementary Fig S1.

In the prior two weeks, 102 persons reported wanting or needing a SARS-CoV-2 test, of whom 62 (61%) were able to get a test, 18 (29%) of whom tested positive. Sixteen (89%) reported subsequent hospitalization (See Supplementary Fig S2). While 18 (29%) reported getting a test the same day they wanted/needed it, about a third (34%) reported waiting 3 or more. Over half (53%) of participants reported waiting 3 or more days to receive a test result; 7 (11 %) had not received their results by the time of the survey.

### Association between prior SARS-CoV-2 infection status, travel history and adoption of NPIs

In unadjusted analysis, self-reported history of SARS-CoV-2 infection was significantly more frequent among those who were younger, male, African American or Hispanic/Latino (p<0.05 for all; Table 2). SARS-CoV-2 infection was also significantly more common among those who reported using public transportation, attending a place of worship, visiting friends or family, attending gatherings of more than 10 people and more than 100 people, and visiting indoor or outdoor venues where people gather (p<0.05 for all; Supplementary Fig S3). Infection was significantly less common in those who reported always practicing social distancing indoors and outdoors (p<0.05 for both).

**Table 2.** Factors associated with ever testing positive for SARS-CoV-2 by univariable and multivariable logistic regression.

| Variable | Ever Tested Positive<br>OR (95% CI) | Ever Tested Positive<br>aOR (95% CI) |
| --- | --- | --- |
| **Race/Ethnicity** |  |  |
| White (ref.) | - | - |
| Black/African American | 3.03 (1.62 – 5.66) | 1.06 (0.44 – 2.54) |
| Hispanic/Latino | 4.14 (1.81 – 9.47) | 2.75 (0.87 – 8.70) |
| **Age** (per 5-year increase) | 0.69 (0.61 – 0.78) | 0.84 (0.71 – 0.99) |
| **Male Gender** | 2.00 (1.13 – 3.45) | 1.55 (0.69 – 3.47) |
| **Annual Household Income** |  |  |
| < \$20,000 (ref.) | - | - |
| \$20,000 – \$39,000 | 0.92 (0.38 – 2.27) | - |
| \$40,000 – \$49,000 | 0.77 (0.26 – 2.25) | - |
| \$50,000 – \$69,000 | 0.48 (0.18 – 1.29) | - |
| \$70,000+ | 0.57 (0.25 – 1.28) | - |
| **Work Outside the Home** | 3.61 (2.08 – 6.29) | - |
| **Practice Social Distancing Indoors** |  |  |
| Never (ref.) | - | - |
| Sometimes | 0.71 (0.31 – 1.63) | 0.26 (0.08 – 0.90) |
| Always | 0.30 (0.14 – 0.67) | 0.32 (0.10 – 0.99) |
| **Practice Social Distancing Outdoors** |  |  |
| Never (ref.) | - | - |
| Sometimes | 0.70 (0.28 – 1.73) | 0.34 (0.10 – 1.19) |
| Always | 0.19 (0.08 – 0.46) | 0.10 (0.03 – 0.33) |
| **Used Public Transport** |  |  |
| Never (ref.) | - | - |
| Once or twice | 8.06 (3.80 – 17.1) | 6.00 (2.13 -16.9) |
| 3 – 7 times | 12.2 (5.64 – 26.3) | 3.80 (1.18 – 12.3) |
| More than 7 times | 18.8 (7.59 – 46.4) | 4.29 (1.12 – 16.5) |
| **Attended Gathering of >10 People** |  |  |
| Never (ref.) | - | - |
| Once or twice | 3.37 (1.56 – 7.25) | - |
| 3 – 7 times | 15.5 (7.22 – 33.4) | - |
| More than 7 times | 28.7 (10.4 – 78.7) | - |
| **Attended Gathering of >100 People** |  |  |
| Never (ref.) | - | - |
| Once or twice | 6.51 (2.77 – 15.3) | - |
| 3 – 7 times | 26.4 (11.37 – 61.4) | - |
| More than 7 times | 34.0 (13.2 – 87.7) | - |
| **Visited Place of Worship** |  |  |
| Never (ref.) | - | - |
| Once or twice | 2.91 (1.06 – 8.02) | 1.41 (0.38 – 5.31) |
| 3 or more times** | 23.9 (12.5 – 45.9) | 16.0 (5.97 – 42.7) |
| **Visited Friends or Family** |  |  |
| Never (ref.) | - | - |
| Once or twice | 1.12 (0.52 – 2.42) | - |
| 3 – 7 times | 3.87 (1.77 – 8.46) | - |
| More than 7 times | 5.42 (1.89 – 15.6) | - |
| **Went to a Grocery Store/Pharmacy** |  |  |
| Never (ref.) | - | - |
| Once or twice | 0.43 (0.17 – 1.13) | - |
| 3 – 7 times | 0.69 (0.27 – 1.81) | - |
| More than 7 times | 2.23 (0.77 – 6.51) | - |
| **Went to an Indoor Bar, Restaurant, Salon or Other Indoor Establishment** |  |  |
| Never (ref.) | - | - |
| Once or twice | 1.10 (0.53 – 2.29) | - |
| 3 – 7 times | 4.14 (1.98 – 8.68) | - |
| More than 7 times | 9.76 (3.83 – 24.9) | - |
| **Went to a Beach, Pool or Other Outdoor Gathering Place** |  |  |
| Never (ref.) | - | - |
| Once or twice | 2.45 (1.16 – 5.18) | - |
| 3 – 7 times | 9.30 (4.47 – 19.3) | - |
| More than 7 times | 8.28 (2.56 – 26.7) | - |
| **Mask Wearing in Public Indoors** |  |  |
| Never or sometimes | - | - |
| Always | 0.63 (0.36 – 1.09) | - |
| **Always Wear Mask in Public Outdoors** |  |  |
| Never or sometimes | - | - |
| Always | 1.06 (0.61 – 1.85) | - |
| **Number of Close Contacts Indoors** |  |  |
| 0 (ref.) | - | - |
| 1 | 3.96 (1.41 – 11.2) | - |
| 2 – 5 | 4.46 (1.92 – 10.3) | - |
| 6 – 10 | 4.87 (1.72 – 13.8) | - |
| >10 | 6.16 (1.72 – 22.0) | - |
| **Number of Close Contacts Outdoors** |  |  |
| 0 (ref.) | - | - |
| 1 | 5.90 (2.00 – 17.4) | - |
| 2 – 5 | 6.30 (2.54 – 15.6) | - |
| 6 – 10 | 9.58 (3.31 – 27.7) | - |
| >10 | 4.26 (0.83 – 21.9) | - |

In multivariable analysis, history of SARS-CoV-2 infection remained significantly more likely among younger respondents, those who took public transportation (adjusted odds ratio [aOR] for 3 or more times vs. none: 4.3; 95% confidence interval [CI]: 1.1 – 16.5), and those who visited a place of worship (aOR for those who visited ≥ 7 times vs. none: 16.0; 95% CI: 6.0 – 42.7), and significantly less common among those who always practiced social distancing (aOR for indoor social distancing: 0.32; 95% CI: 0.10 – 0.98; aOR for outdoor social distancing: 0.10; 95% CI: 0.03 – 0.33) (Table 2). Associations were similar in sensitivity analyses of self-reported SARS-CoV-2 infection in the prior 2 weeks (Supplementary Table S4).

## Discussion

The COVID-19 Pandemic Pulse Study utilized an online panel survey methodology to provide a rapid, cost-efficient snapshot of travel patterns and adoption of NPI across population subgroups in Maryland. These data indicate that non-essential travel and uneven adoption of NPI could potentially influence community transmission of SARS-CoV-2 infection in Maryland. Over two thirds of the respondents reported recent travel for non-essential services; self-reported SARS-CoV-2 infection was significantly more common among those who reported using public transport or visiting places of worship and significantly less common among those whom always practiced social distancing. Significant differences in the adoption of NPI were observed by age, race/ethnicity and income suggesting that communication campaigns tailor messaging to specific subgroups.

Strong, positive dose-response relationships were demonstrated between recent movement and likelihood of being diagnosed with SARS-CoV-2 infection. The more frequently an individual participated in an activity, the more likely they were to have tested positive. Additionally, consistent with ecologic analyses^8^, these data suggest lower levels of SARS-CoV-2 positivity among those always practicing social distancing. Indeed, when adjusting for social distancing, most associations between recent movement and SARS-CoV-2 positivity were no longer statistically significant. This supports public health messaging that incorporating appropriate NPI while visiting indoor and outdoor venues helps reduce SARS-CoV-2 transmission. Importantly, this was not the case with all forms of movement; using public transport and visiting a place of worship in the prior 2 weeks were significantly associated with SARS-CoV-2 positivity even after adjusting for social distancing. Of course, while mask use is mandated for public transport, social distancing may be challenging and use may reflect necessity vs. personal choice. While in Maryland, many religious gatherings moved to remote services, indoor religious gatherings were allowable in Maryland with restrictions at the time of the survey, We specifically surveyed about physically visiting places of worship but were unable to discriminate the reasons why persons attended a place of worship (e.g., service vs. food distribution, Narcotics Anonymous, etc.). It is important to note that data on movement and NPI adoption, albeit self-reported, was limited to the prior two-week timeframe to minimize potential recall bias. Notably, objective measures of movement such as cell phone data analytics cannot easily discriminate between activities, venues, or practice of NPI during mobility.

While these data demonstrated a negative association between consistent indoor mask use and ever testing positive for SARS-CoV-2, this association failed to achieve statistical significance. Collection of data related to mask use is nuanced as there are several factors that can affect the efficacy of masks that are challenging to collect via this online format such as fit, type of mask, frequency of touching/adjusting mask, etc. In sensitivity analyses that restricted analyses to recent mask use and recent SARS-CoV-2 infection, consistent indoor mask use was significantly associated with a lower likelihood of infection.

These analyses could not establish temporality between the exposures and SARS-CoV-2 positivity and may reflect that those who tested positive were more likely to practice these behaviors when they were infected or that they changed their behaviors after testing positive potentially because of altered risk perception. Sensitivity analyses demonstrating similar associations with recent SARS-CoV-2 infection and the strong dose-response associations observed suggest that that reverse causality does not likely explain the associations observed. Regardless of directionality, these findings have implications for community transmission risk particularly in light of incomplete understanding of viral shedding and infectiousness among infected persons and the role and duration of acquired immunity on protection from SARS-CoV-2 reinfection.^22-24^ Of interest, these data were collected just as Maryland began to relax restrictions and the state is now, approximately one month later, witnessing an uptick^16^ in cases particularly among younger persons, the strata who reported maximum mobility in this survey.

Aside from the widescale adoption of NPI, early diagnosis, appropriate contact tracing measures, and isolation and/or referral to care are critical to curtailing SARS-CoV-2 transmission.^25^ It is vital that all persons who seek a test are able to obtain one with minimal delays followed by prompt receipt of results. In this sample, almost 40 percent of those who wanted a test were not able to get one; furthermore, even in those tested, there were significant delays in being able to obtain a test, and over half waited three or more days for results. To effectively curb transmission, these delays need to be addressed improving access and timely result reporting.

This sample of 1,030 respondents was recruited in under two weeks at a cost of $3,000. While this sample cannot necessarily be considered fully representative of Maryland, several advantages of the approach are worth highlighting. First, in a rapidly evolving epidemic where behaviors and practices are constantly changing, this approach did not require any face-to-face visits maximizing participant safety and minimizing survey administration costs. Second, while there is bias in that individuals require internet connectivity to participate, internet access has been improving and has been facilitated by recent discounts offered by major providers. Indeed, it is estimated that 86% of Maryland residents have internet connectivity.^19^ Despite this, our sample likely misses homeless populations and very low-income groups, two populations where NPI adoption may be especially challenging.^20^ Among those with internet connectivity, it is not possible to sample randomly and the “frame error” of those willing to participate in such panels is likely large but cannot be proven to be systematic. Third, utilizing quotas for various key demographic characteristics, online samples can be structured to recruit samples with demographic distributions comparable to the target population.^21^ Notably, although this sample was not intended to estimate SARS-CoV-2 prevalence, the self-reported positivity rates reflect case count data in Maryland^16^ with higher positivity among men, Hispanic, African-American, and younger populations. Finally, any biases will likely remain constant over time allowing for examining trends longitudinally. To monitor these trends, we developed an online interactive dashboard (<u>sclipman.github.io/PandemicPulse</u>).

Limitations notwithstanding, we present a rapid cost-efficient approach of monitoring NPI adoption and adherence which can help inform public health response. While our survey illustrated this approach within a single state, the rapidity and efficiency of this methodology can be easily replicated in other settings recognizing the highly variable and geographically localized SARS-CoV-2 transmission patterns and risk mitigation responses. Repeating these surveys over time in a given population can unveil additional insights around changes in population behaviors potentially informing adaptive responses to evolving disease dynamics. Overall, these data continue to highlight the role of movement and social distancing on SARS-CoV-2 transmission risk. In Maryland, these data support targeted COVID-19 messaging to young adults given high rates of positivity as well as the lower rates of adoption of NPIs; establishing partnerships with faith-based organizations could also be critical to curbing the spread. Moreover, measures need to be implemented to make public transportation safe for those who need to use it and to improve access to SARS-CoV-2 testing. Continued monitoring of the adoption of NPIs, access to testing and the subsequent impact on SARS-CoV-2 transmission in Maryland as well as more broadly will be critical for pandemic control.

## Data Availability

The data that support the findings of this study are available from the corresponding author upon reasonable request. Aggregate data is available online through the Pandemic Pulse dashboard.

https://sclipman.github.io/PandemicPulse/

## Acknowledgements

We would like to gratefully acknowledge the contributions of Mr. Adebola Adegbesan who worked closely with our team in the recruitment of the study sample, as well as the contributions of Ms. Hannah Manley, who assisted with data cleaning. This work was supported by the Johns Hopkins COVID-19 Research Response Program. APW is funded by a Career Award at the Scientific Interface by the Burroughs Wellcome Fund and by the National Library of Medicine of the National Institutes of Health under Award Number DP2LM013102. SSS is funded by DP2DA040244. The content is solely the responsibility of the authors and does not necessarily represent the official views of the Johns Hopkins University or the National Institutes of Health.

## Supplemental Material

**Supplemental Fig S1.**
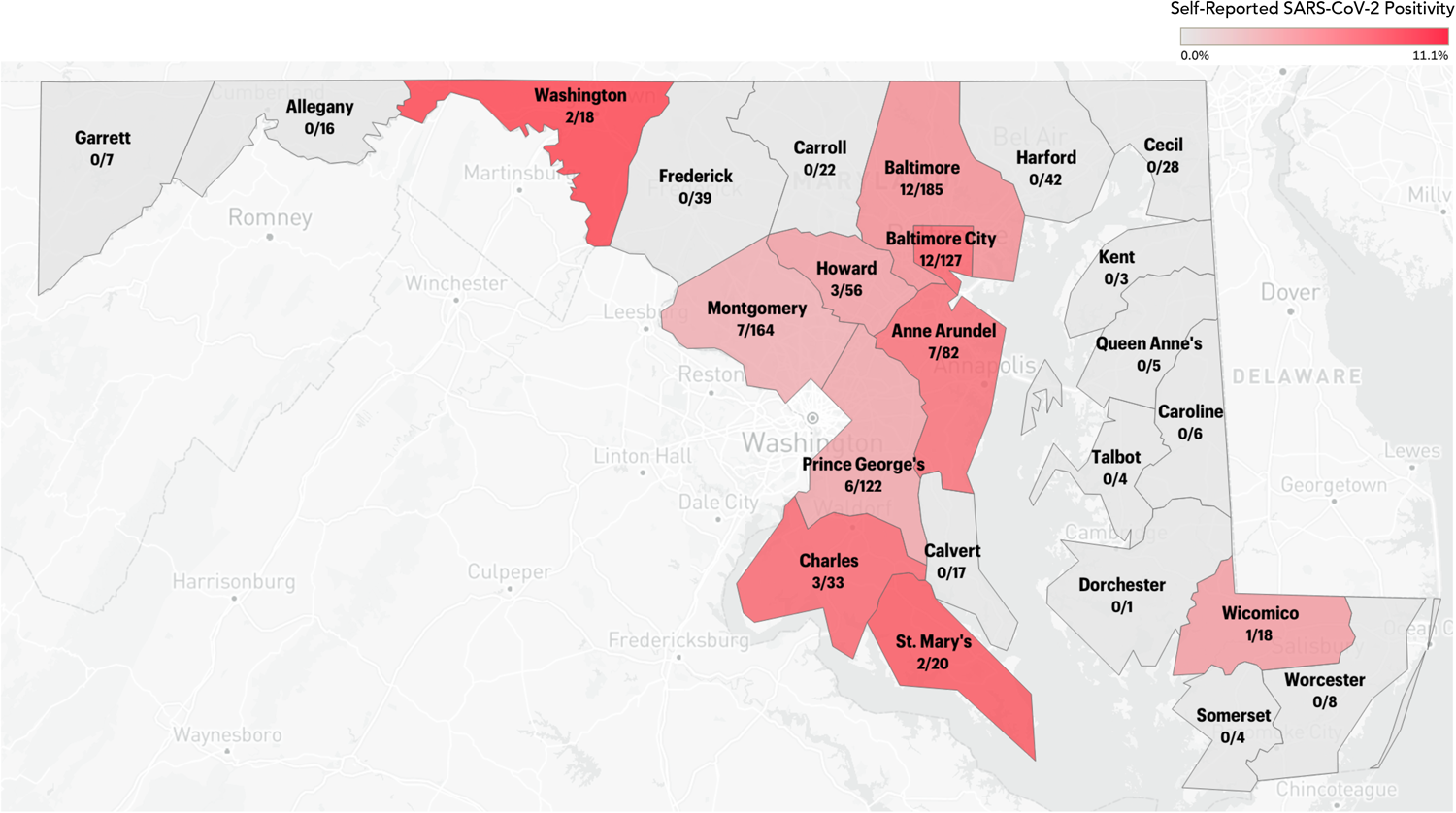
Self-reported SARS-CoV-2 positivity by Maryland county. Numbers below county labels represent the number of positives over the total number of respondents from that county.

**Supplemental Fig S2.**
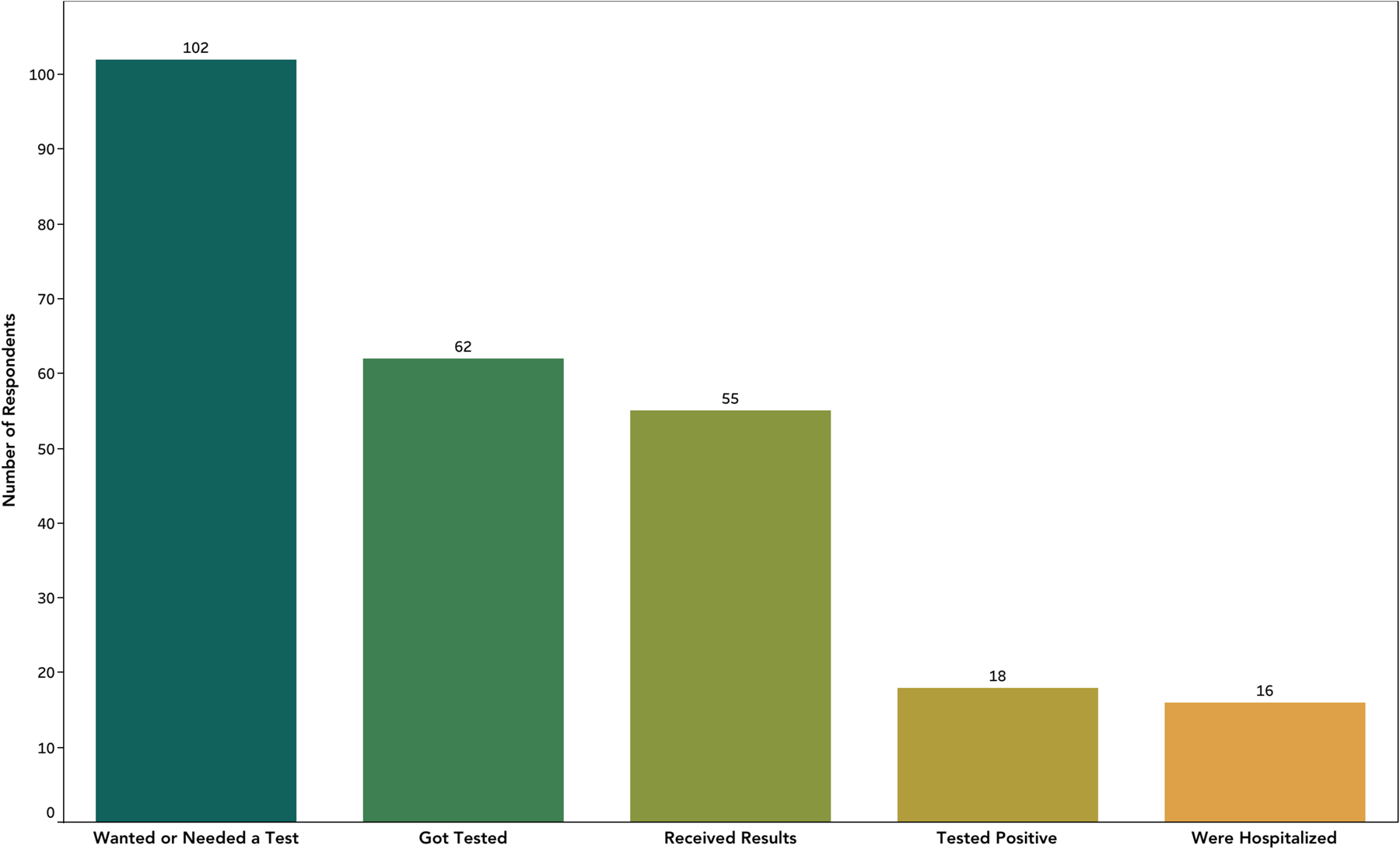
SARS-CoV-2 testing cascade. All numbers reflect behaviors/results in 2 weeks.

**Supplemental Fig S3.**
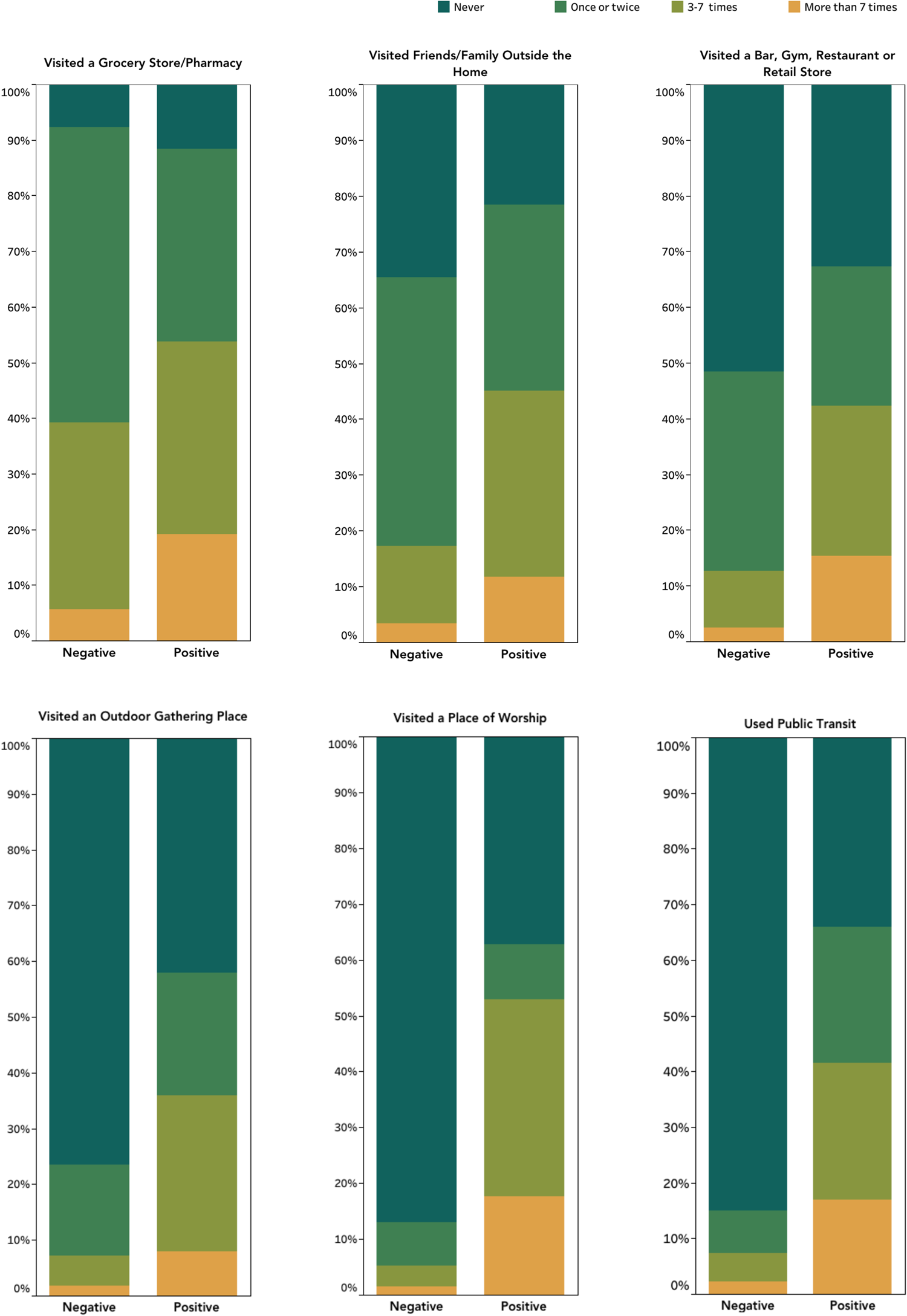
Participant mobility patterns in the prior 2 weeks by SARS-CoV-2 infection status.

**Supplemental Table S4.**
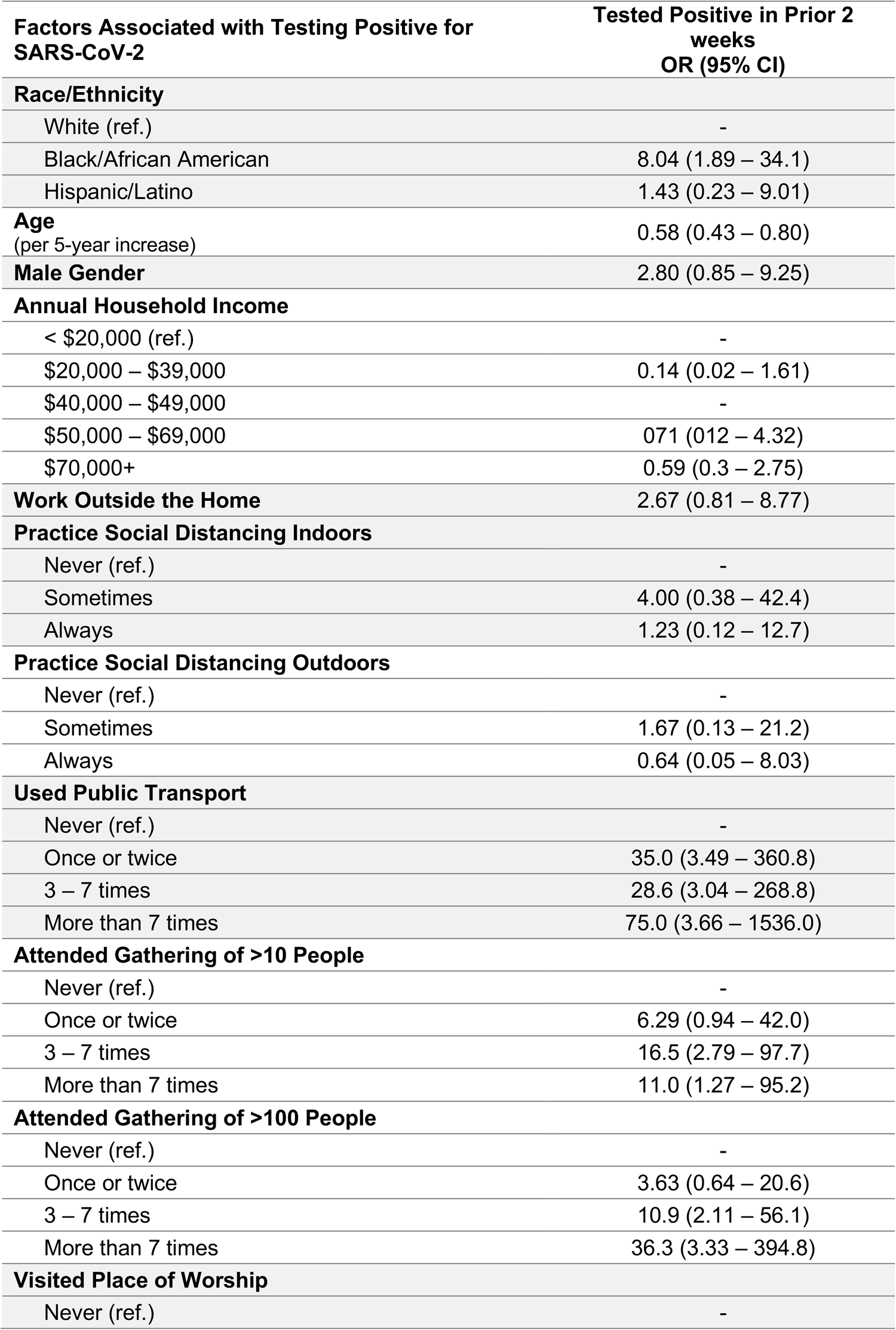
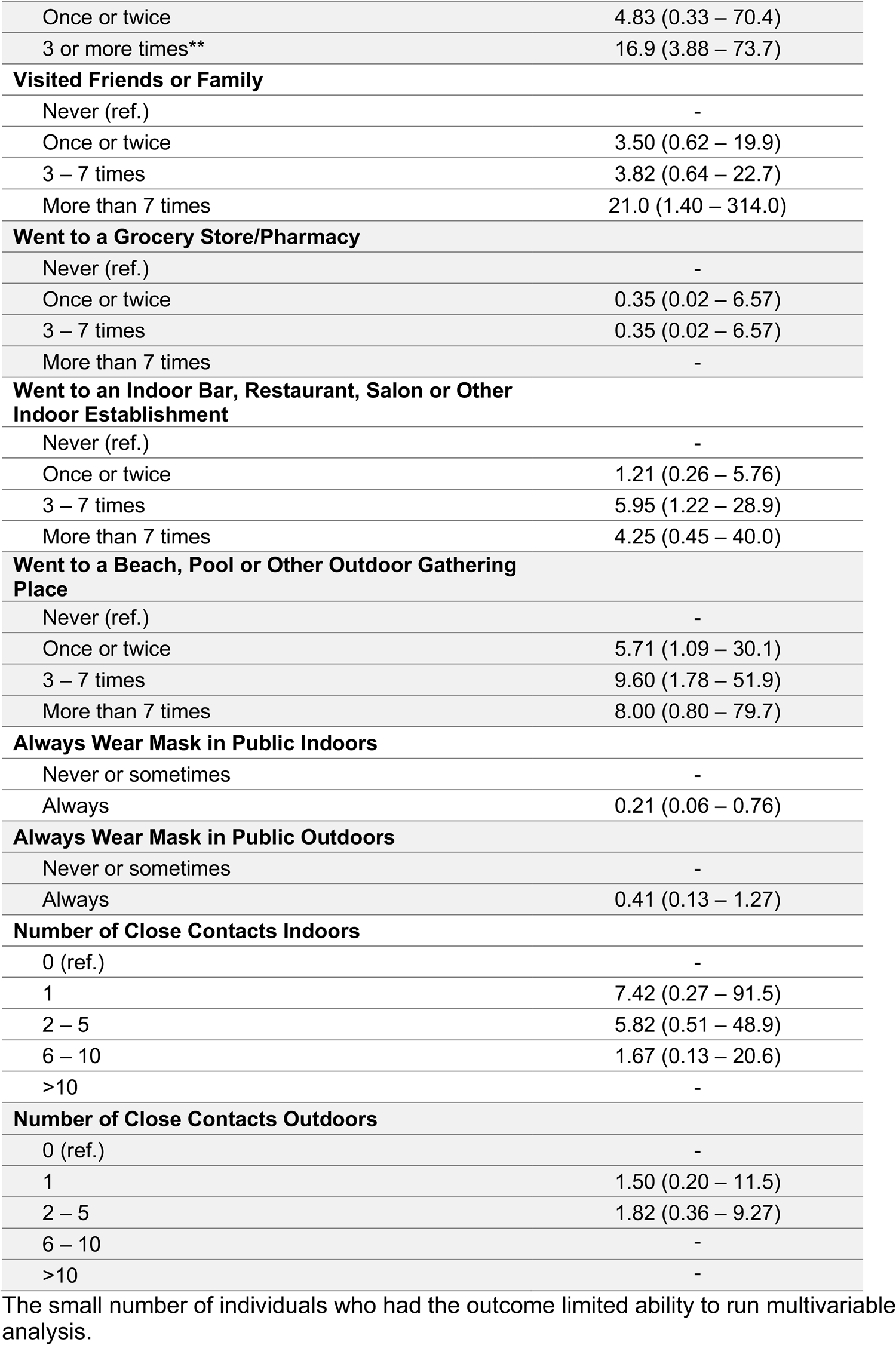
Factors associated with testing positive for SARS-CoV-2 in prior two weeks by univariable logistic regression.

| **Factors Associated with Testing Positive for SARS-CoV-2** | **Tested Positive in Prior 2 weeks**<br>**OR (95% CI)** |
| --- | --- |
| **Race/Ethnicity** |  |
| White (ref.) | - |
| Black/African American | 8.04 (1.89 – 34.1) |
| Hispanic/Latino | 1.43 (0.23 – 9.01) |
| **Age**<br>(per 5-year increase) | 0.58 (0.43 – 0.80) |
| **Male Gender** | 2.80 (0.85 – 9.25) |
| **Annual Household Income** |  |
| < \$20,000 (ref.) | - |
| \$20,000 – \$39,000 | 0.14 (0.02 – 1.61) |
| \$40,000 – \$49,000 | - |
| \$50,000 – \$69,000 | 0.71 (0.12 – 4.32) |
| \$70,000+ | 0.59 (0.3 – 2.75) |
| **Work Outside the Home** | 2.67 (0.81 – 8.77) |
| **Practice Social Distancing Indoors** |  |
| Never (ref.) | - |
| Sometimes | 4.00 (0.38 – 42.4) |
| Always | 1.23 (0.12 – 12.7) |
| **Practice Social Distancing Outdoors** |  |
| Never (ref.) | - |
| Sometimes | 1.67 (0.13 – 21.2) |
| Always | 0.64 (0.05 – 8.03) |
| **Used Public Transport** |  |
| Never (ref.) | - |
| Once or twice | 35.0 (3.49 – 360.8) |
| 3 – 7 times | 28.6 (3.04 – 268.8) |
| More than 7 times | 75.0 (3.66 – 1536.0) |
| **Attended Gathering of >10 People** |  |
| Never (ref.) | - |
| Once or twice | 6.29 (0.94 – 42.0) |
| 3 – 7 times | 16.5 (2.79 – 97.7) |
| More than 7 times | 11.0 (1.27 – 95.2) |
| **Attended Gathering of >100 People** |  |
| Never (ref.) | - |
| Once or twice | 3.63 (0.64 – 20.6) |
| 3 – 7 times | 10.9 (2.11 – 56.1) |
| More than 7 times | 36.3 (3.33 – 394.8) |
| **Visited Place of Worship** |  |
| Never (ref.) | - |
| Once or twice | 4.83 (0.33 – 70.4) |
| 3 or more times** | 16.9 (3.88 – 73.7) |
| **Visited Friends or Family** |  |
| Never (ref.) | - |
| Once or twice | 3.50 (0.62 – 19.9) |
| 3 – 7 times | 3.82 (0.64 – 22.7) |
| More than 7 times | 21.0 (1.40 – 314.0) |
| **Went to a Grocery Store/Pharmacy** |  |
| Never (ref.) | - |
| Once or twice | 0.35 (0.02 – 6.57) |
| 3 – 7 times | 0.35 (0.02 – 6.57) |
| More than 7 times | - |
| **Went to an Indoor Bar, Restaurant, Salon or Other Indoor Establishment** |  |
| Never (ref.) | - |
| Once or twice | 1.21 (0.26 – 5.76) |
| 3 – 7 times | 5.95 (1.22 – 28.9) |
| More than 7 times | 4.25 (0.45 – 40.0) |
| **Went to a Beach, Pool or Other Outdoor Gathering Place** |  |
| Never (ref.) | - |
| Once or twice | 5.71 (1.09 – 30.1) |
| 3 – 7 times | 9.60 (1.78 – 51.9) |
| More than 7 times | 8.00 (0.80 – 79.7) |
| **Always Wear Mask in Public Indoors** |  |
| Never or sometimes | - |
| Always | 0.21 (0.06 – 0.76) |
| **Always Wear Mask in Public Outdoors** |  |
| Never or sometimes | - |
| Always | 0.41 (0.13 – 1.27) |
| **Number of Close Contacts Indoors** |  |
| 0 (ref.) | - |
| 1 | 7.42 (0.27 – 91.5) |
| 2 – 5 | 5.82 (0.51 – 48.9) |
| 6 – 10 | 1.67 (0.13 – 20.6) |
| >10 | - |
| **Number of Close Contacts Outdoors** |  |
| 0 (ref.) | - |
| 1 | 1.50 (0.20 – 11.5) |
| 2 – 5 | 1.82 (0.36 – 9.27) |
| 6 – 10 | - |
| >10 | - |

The small number of individuals who had the outcome limited ability to run multivariable analysis.

